# Clinical picture and diagnosis of comorbidity of disseminated pulmonary tuberculosis, coronavirus, pneumocystis and pneumococcal pneumonia in patients with advanced HIV infection

**DOI:** 10.1101/2025.01.11.24319602

**Authors:** A. V. Mishina, V. Yu. Mishin, I. V. Shashenkov

**Affiliations:** FSBEI HE Russian University of Medicine of the Ministry of Health of Russia, Moscow, Russia; FBHI The Tuberculosis Clinical Hospital N□3 named after Professor G.A. Zakharyin, Moscow, Russia

## Abstract

**Purpose:** The aim of the study was to evaluate the features of clinic and diagnosis of comorbidity of pulmonary tuberculosis, coronavirus (CVP), pneumocystis (PCP) and pneumococcal (PCcP) pneumonia in patients in the late stages of HIV infection with immunodeficiency (ID).

**Materials and methods:** The prospective study included 120 newly diagnosed patients with disseminated pulmonary tuberculosis with isolation of *Mycobacterium tuberculosis* and IVB stage of HIV infection in the progression phase and in the absence of antiretroviral therapy aged 29-53 years, who were randomized into the main 1A and 2A groups and the 1B and 2B groups of comparison. The 1A group included 29 patients with comorbidity of disseminated pulmonary tuberculosis, coronavirus and pneumocystis pneumonia, in 2A - 31 patients with comorbidity of disseminated pulmonary tuberculosis, CVP and PCcP, while the 1B and 2B groups included 29 and 31 similar patients, but without CVP. PCR for SARS-CoV-2 RNA was used in naso- and oropharyngeal swabs, in sputum or in endotracheal aspirate to diagnose CVP. For the detection of *Pneumocystis jirovecii*, the causative agent of PCP, a microscopic examination of diagnostic material from the respiratory tract was carried out with staining according to Romanowsky-Giemsa and according to Grocott-Gomori, while for detection of *Streptococcus pneumoniae*, the causative agent of pneumococcal pneumonia, the diagnostic material was sown on special nutrient media, with the determination of drug resistance of the obtained culture to broad-spectrum antibiotics. Statistical data processing was carried out using Microsoft Office Excel 2019 with calculation of mean value of the indicator in the group, standard error and confidence interval.

**Results:** The comorbidity of disseminated pulmonary tuberculosis, CVP, PCP and PCcP in patients in the late stages of HIV infection in the phase of progression and in the absence of antiretroviral therapy were characterized by severe immunodeficiency, generalization of tuberculosis with multiple extrapulmonary lesions and severe pneumonia. This determines the similarity of clinical manifestations and respiratory symptoms as well as also makes it difficult to visualize computed tomographic changes consisting of complex of simultaneous combination of four pathological syndromes: dissemination, pleural pathology, increased pulmonary pattern and adenopathy. Simultaneous overlap of several pathologies with the same type of clinical manifestations and computed tomographic changes requires a complex etiological diagnosis of the specific diseases to prescribe timely complex treatment and reduce mortality among that severe contingent of patients.

**Conclusion:** Patients with disseminated pulmonary tuberculosis and HIV infection represent a high-risk group for COVID-19 infection and development of CVP (and in in cases of severe immunodeficiency - PCP and PCcP) - they should be regularly subjected to preventive examinations for timely detection of COVID-19, CVP, PCP and PCcP and for purpose of their emergency hospitalization and timely treatment.

## Introduction

The problem of comorbid infectious pathology of the respiratory system among HIV-infected patients is becoming increasingly urgent, especially in the late stages of HIV with severe immunodeficiency (ID), when secondary opportunistic lung infections (OLI) develop, occurring with damage to the lower respiratory tract, the causative agents of which are bacteria and less often fungi and viruses (1,2). The most common secondary bacterial disease in HIV-infected persons with ID is tuberculosis, most commonly occurring as generalized disseminated pulmonary tuberculosis (DPT) with multiple extrapulmonary foci. Other frequent OLIs are PCP caused by *Pneumocystis jirovecii* and PCcP caused by *Streptococcus pneumoniae*, that provoke an increase in the respiratory symptoms and development of bilateral polysegmental pathological changes in lung tissue and interstitial changes of the “ground glass” type which significantly complicates diagnosis of a comorbid disease and is one of the main reasons for hospitalization of patients with HIV infection and the most common cause of their death, especially in cases of comorbidity with tuberculosis.

The SARS-CoV-2 has created serious clinical problems in patients with HIV infection due to development of multiple organ failure, septic shock and venous thromboembolism with severe damage to the respiratory system and CVP, that is especially dangerous in HIV infection with ID and OLI and makes such patients especially dangerous in terms of infection for healthy population (3, 4, 5, 6, 7, 8, 9).

### Purpose

**The purpose of the study** was to evaluate the features of clinical manifestations and diagnosis of comorbidity of DPT, CVP, PCP and PCcP in patients in the late stages of HIV infection with ID.

## Materials and methods

The prospective study included 120 newly diagnosed patients with DPT with isolation of M. *tuberculosis*, with IVB stage of HIV infection in the progression phase and in the absence of antiretroviral therapy (ART), aged 29-53 years. Ther were 77 men (64.2%) and 43 women (35.8%). Patients were randomized into the main 1A and 1B groups and the 2A and 2B groups of comparison.

The group 1A was composed of 29 patients having been diagnosed with comorbidity of DPT, CVP and PCP; and group 1B of 31 patients diagnosed with comorbidity of DPT, CVP and PCcP. The comparison groups 2A and 2B were composed of 29 and 31 patients, respectively, who were not diagnosed with CVP. They were selected according to the “copy-pair” principle in relation to patients of the main groups, they also were almost identical in social, age, sexual, clinical parameters, stage of HIV infection, severity of ID and absence of other OLIs having been developed in the later stages of HIV.

For etiological diagnosis of DPT, in the diagnostic material of the respiratory tract (sputum, bronchoalveolar lavage, biopsy material obtained from bronchoscopy and punctures of the intrathoracic lymph nodes) and other organs (blood, urine, feces and peripheral lymph node punctures) fluorescent microscopy and PCR of *M. tuberculosis* DNA were performed, sowing on a dense Löwenstein-Jensen culture medium and on a liquid culture in the BACTEC MGIT 960 automated system, with the determination of drug resistance of the obtained culture *of M. tuberculosis* to anti-tuberculosis drugs (ATD) using the method of absolute concentrations.

The SARS-CoV-2 RNA PCR was used in naso- and oropharyngeal swabs, sputum, or endotracheal aspirate to diagnose COVID-19 and CVP. To detect *P. jirovecii*, the causative agent of PCP, in oropharyngeal swabs and in bronchoalveolar lavage, microscopic examination was used when staining according to Romanovsky-Giemsa and according to Grocott-Gomori and immunofluorescence reaction with mono- and polyclonal antibodies and determining the level of immunoglobulins of class G and M. The high level of total activity of lactate dehydrogenase and a decrease in oxygen saturation in the blood were taken into account, and for etiological diagnosis of PCcP caused by S. *pneumoniae*, the diagnostic material of the respiratory tract was examined by microscopy of smears stained by Gram and Gis method and the diagnostic material was sown on special nutrient media (blood agar and serum broth, 10% bile broth), with determination of drug resistance of the obtained culture to broad-spectrum antibiotics (BSA) by disco-diffusion method or by serial dilution.

To exclude other OLI pathogens such as mycobacteriosis of the lungs caused by *Mycobacterium nontuberculosis*, hospital-acquired bacterial pneumonia - *Staphylococcus aureus* and *Haemophilus influenzae*, atypical pneumonia - *Legionella pneumophila, Mycoplasma pneumoniae* and *Chlamydophila pneumoniae*, candidal pneumonia - *Candida albicans* and viral pneumonia - *Herpes simplex virus-1* or *Cytomegalovirus Human*, microbiological, virological, immunological methods and PCR of the diagnostic material from respiratory tract, blood, urine and cerebrospinal fluid were used.

All patients underwent comprehensive clinical, laboratory, immunological (determination of number of CD4+ lymphocytes by flow cytofluorimetry and viral load by number of copies of HIV RNA in the peripheral blood) and radiation examination, which included computed (CT), magnetic resonance imaging and ultrasound of the chest organs (ChO) and internal organs.

Statistical data processing was carried out using the Microsoft Office Excel 2019 program, with calculation of means of indicator in the group, standard error of mean and confidence interval. The *p-test* was determined from the Student’s table. Differences between arithmetic values of means were considered significant at *p* <0.05.

## Results

In all 120 patients from all groups, duration of HIV infection was 8-12 years. All of them were registered with an AIDS center, which was practically not visited due to social maladjustment and lack of adherence to examination and treatment. All the patients were addicted to drugs, consumed alcoholic beverages and were tobacco smokers. All patients were diagnosed with viral hepatitis B or C and 79 of them (65,8±4,3%) with chronic obstructive pulmonary disease (COPD).

All the 120 patients had symptoms of acute inflammatory respiratory disease and bilateral inflammatory lung lesions when they came to primary health care facilities where they were suspected of having pulmonary tuberculosis. They were sent to the TD. There, a comprehensive microbiological study of the diagnostic material of the respiratory tract has revealed *M. tuberculosis* and CT of the ChO and visualized bilateral lung damage typical for DPT. The patients were hospitalized. In the admission unit, 60 patients in the 1A and 1B groups were diagnosed as having COVID-19 and CVP, they were isolated in the observational unit (“red zone”). The 60 patients of 2A and 2B were COVID-19-free and were referred to the TB and HIV unit.

In patients of 1A and 2A groups, microscopic examination and immunofluorescence reactions with mono- and polyclonal antibodies of diagnostic material of the respiratory tract revealed *P. jirovecii*, a diagnosis of PCP was established, while in patients of 1B and 2B groups when sowing diagnostic material from the respiratory tract to special nutrient media, a culture *of S. pneumoniae* was obtained and a diagnosis of PCcP was established. The distribution of patients in the observed groups by the number of CD4+ lymphocytes in 1 μL of blood is presented in Table 1.

**Table 1.** Distribution of patients in the observed groups by CD4+ lymphocyte in 1 μL of blood (M±m)

| Table 1. Distribution of patients in the observed groups by CD4+ lymphocyte in 1 μL of blood (M±m) | | | | | |
| --- | --- | --- | --- | --- | --- |
| Groups for examination | Number of patients, abs. (%) | Lymphocyte count |  |  |  |
|  |  | 50–30 | 29–20 | 19–10 | less than 9 |
| 1A | 29 (100) | 6 [20.7±7.2] | 7 [24.1±7.9] | 11 [37.9±9.0] | 5 [17.2±7.0] |
| 2A | 31 (100) | 7 [22.6±7.5] | 7 [22.6±7.5] | 13 [41.9±8.8] | 4 [12.9±6.0] |
| 1B | 29 (100) | 5 [17.2±7.0] | 8 [27.6±8.3] | 10 [34.5±8.8] | 6 [20.7±7.2] |
| 2B | 31 (100) | 8 [25.8±7.8] | 5 [16.1±6.6] | 15 [48.4±9.0] | 6 [20.7±7.2] |

As it is shown in Table 1, in all groups the number of CD4+ lymphocytes did not exceed 50 in 1 μL of blood and their quantitative ratio for the observed groups practically did not differ from each other.

In the 1A group, 20.7% of patients had CD4+ lymphocyte count in the range of 50-30 cells/μL of blood: 24.1% had 20-29, 37.9% had 19-10 and 17.2% had less than 9, and in the 1B group, there were 17.2, 27.6, 34.5 and 20.7%, respectively (p> 0.05). In the 2A group, 22.6% of patients had CD4+ lymphocyte count in the range of 50-30 cells/μL of blood, 22.6% had 20-29, 41.9% had 19-10 and 12.9% had less than 9, and in the 2B group, there were 25.8, 16.1, 48.4 and 9.7%, respectively (p> 0.05).

The mean number of CD4+ lymphocytes in 1 μL of blood was also similar, in groups 1A and 2B there were 24,8±0,37 and 26,5±0,43 cells/μL, respectively, and in groups 2A and 2B - 20,3±0,45 and 28,3±0,33 (p> 0.05), respectively, At the same time, the viral load in patients of the observed groups was more than 500 thousand RNA HIV copies per 1 mL of blood.

Thus, in patients of 1A group with comorbidity of DPT, CVP and PCP, 2A group with comorbidity of DPT and PCP and without CVP, 1B group with comorbidity of DPT, CVP and PCcP and 2B group with comorbidity of DPT and PCcP and without CVP, with the IVB stage of HIV infection in the progression phase and in the absence of ART, a significant decrease in number of CD4+ lymphocytes was noted (from 50 cells per 1 μL of blood and lower) with an mean CD4+ cell count not exceeding 30 per 1 μL of blood and high viral load of more than 500 thousand of RNA HIV copies per 1 mL of blood, which indicates a pronounced ID. This demonstrates the same type of low immune response to simultaneous exposure to *M. tuberculos*is, *P. jirovecii and S. pneumoni*ae and determines prevalence of pathological lung lesions and the similar type of clinical manifestations of comorbid disease, but we have not established the effect of COVID-19 and CVP on reducing the number of CD4+ lymphocytes and the severity of HIV viral load.

In patients of the examined groups, DPT was combined with tuberculosis generalization with multiple extrapulmonary specific lesions of various organs confirmed by isolation of *M. tuberculosis* from diagnostic material from various organs. In patients of 1A group, two organs were affected in 10 patients, three - in 6, four - in 3 and five - in 2, and in patient s of 2A group two organs - in 10, three - in 7, four - in 2 and five - in 1. A similar frequency of extrapulmonary lesions was recorded in groups 1B and 2B, where two organs were affected in 13 and 7 patients, three - in 5 and 6, four - in 2 and 5, five - in 2. The most frequent extrapulmonary localizations in patients in the observed groups were intrathoracic, peripheral and mesenteric lymph nodes, intestines, genitourinary system and central nervous system. Lesions of the spleen, bones and joints were slightly less common, and lesions of the thyroid gland, adrenal glands, pericardium, inner ear - in single patients.

Therefore, in patients of 1A group with comorbidity of DPT, CVP and PCP, 2A group with comorbidity of DPT and PCP and without CVP, 1B group with comorbidity of DPT, CVP and PCP and 2B group with comorbidity of DPT and PCcP and without CVP, with the IVB stage of HIV infection, severe ID in the phase of progression and in the absence of ART, the multi-organ pathology was characterized by generalized pattern of tuberculosis that reflects severity of the clinical manifestations of the comorbid disease.

The clinical pattern of the disease in patients in the observed groups practically did not differ between each other and was characterized by a pronounced intoxication syndrome and general inflammatory changes with weight loss, headache, myalgia, neuropathy, encephalopathy, palpitations, pale skin, fever, chills, decreased SpO2 and increasing pulmonary heart failure, i.e. essentially the pattern of development of septic shock. All this was also combined with symptoms of damage to other organs and systems. In laboratory tests, the indicators of inflammation typical for septic state were determined: a high leukocytosis was recorded in the blood test - more than 1510^9^/L×, lymphopenia - less than 1.210^9^/L and a sharply accelerated erythrocyte sedimentation rate - more than 40 mm/h, and in biochemical study - high level of C-reactive protein> 100 mg/L. The pattern of inflammatory changes in the respiratory system in all patients also did not differ between them significantly and was characterized by shortness of breath, cough, bronchospasm, secretion of mucopurulent sputum and the presence of diverse wheezing in the lungs.

In the patients from groups 1A and 1B, there were symptoms typical for COVID-19: anosmia, dysgeusia and sensorineural hearing loss, hypoxemia, disseminated intravascular coagulation, thrombosis or thromboembolism and in some cases the Guillain-Barré syndrome. However, similar clinical manifestations occurred with varying frequency in patients of groups 2A and 2B without COVID-19, which probably determined by late stage of HIV infection with severe ID and severe comorbid pathology, that was significantly associated not only with generalized tuberculosis, but also with severity of course of CVP, PCP and PCcP.

Thus, in patients from 1A group with comorbidity of DPT, CVP and PCP, 2A group with comorbidity of DPT and PCP and without CVP, 1B group with comorbidity of DPT, CVP and PCcP and 2B with comorbidity of DPT and PCcP and without CVP, with IVB stage HIV infection, marked by ID, in the progression phase and in the absence of ART clinical presentation, characterized by intoxication syndrome, general inflammatory and respiratory manifestations, was almost the same and nonspecific, and features of clinical course of comorbid disease indicated the similarity of the clinical manifestations and course of DPT, CVP, PCP and PCcP.

During CT of the ChO, a complex of simultaneous combination of 4 pathological syndromes was visualized: dissemination, pleural pathology, increased pulmonary pattern and adenopathy. The dissemination syndrome was represented by foci of different sizes (from small to large) and intensity (from low to high), with a tendency to merge and form infiltrates of a non-homogeneous nature, with destruction of lung tissue and bronchogenic contamination. Pleural involvement syndrome was manifested by compaction of the interlobular and parietal pleura, and in more than 50% of patients - with development of exudative pleurisy or pleural empyema. The syndrome of strengthening and deformation of the pulmonary pattern had a “mesh” character due to development of interstitial pneumonia in lymphohematogenic dissemination with diffuse decrease in the transparency of the lung tissue, the development of cystic-dystrophic changes and areas of consolidation like “ground glass.” Adenopathy syndrome was represented by bilateral enlargement of the intrathoracic lymph nodes with infiltrative changes along the periphery.

At the same time, infiltrates were more often visualized in patients of groups 2A and 2B, against background of which bronchial lumens were detected, mainly in the lower lobes of the lungs, with formation of small abscesses, the appearance of effusion in the pleural cavity, but these changes also occurred in patients from groups 1A and 1B. There was an overlap of several pathologies and changes developing in the lungs in late stages of HIV infection with ID including those directly related to HIV infection itself in the form of lymphoid interstitial pneumonia, nonspecific interstitial pneumonia, pulmonary hypertension and COPD with emphysema. At the same time, the volume of lung damage in all patients was 80-100% and was almost comparable. It was not possible to differentiate these changes visible during CT of the ChO by specific pathologies due to similarity of the CT signs, while diagnosis was possible only with microbiological, virological and molecular genetic studies of the specific pathogens.

Thus, in patients of 1A group with comorbidity of DPT, CVP and PCP, group 2A with comorbidity of DPT and PCP and without CVP, group 1B with comorbidity of DPT, CVP and PCcP and group 2B with comorbidity of DPT and PCcP and without CVP, with the IVB stage of HIV infection, pronounced ID, in the phase of progression and in the absence of ART, the clinical picture was characterized by intoxication syndrome, general inflammatory and respiratory manifestations, almost the same and nonspecific, and features of clinical course of comorbid disease could be determined only when a specific pathogen was identified. As examples, the CT images of the ChO of patients with IVB stage of HIV infection with ID in the phase of progression and in absence of ART are presented below that illustrate the similarity of visualization of pathological change all groups (Fig. 1, 2).

**Fig. 1.**
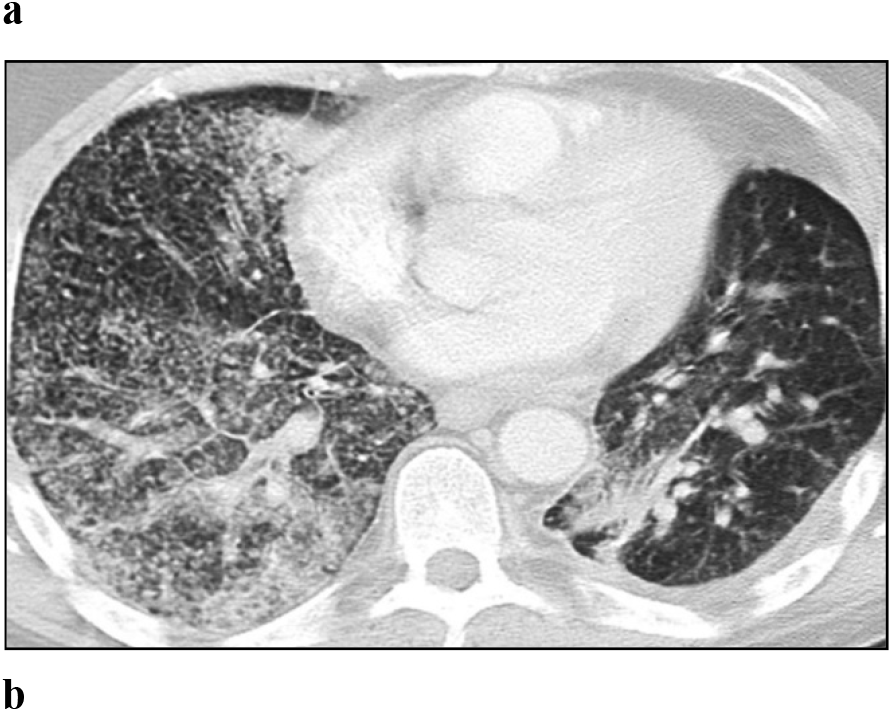
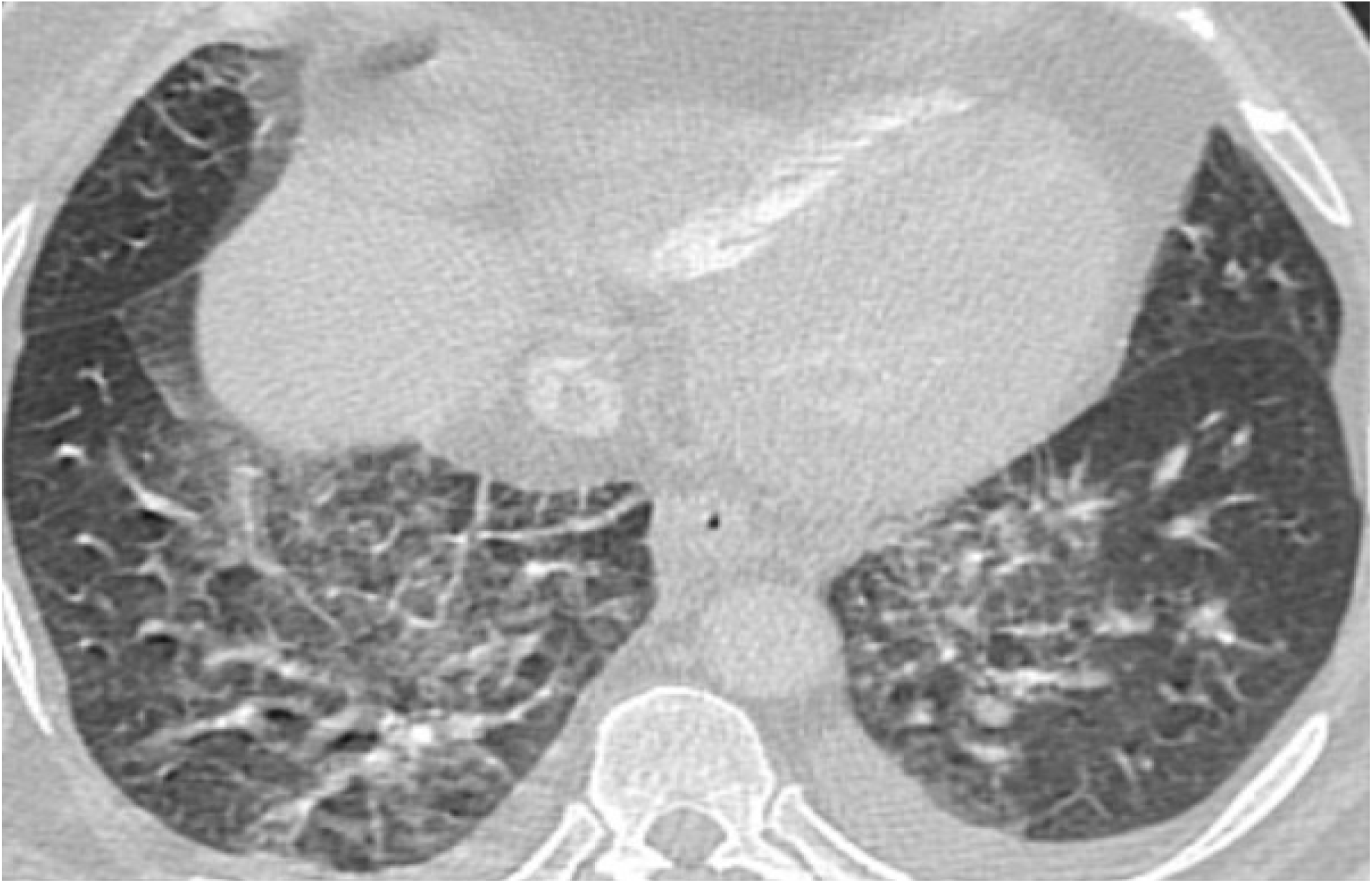
The CT of the ChO. Axial projection, pulmonary window mode: a - Patient with stage IVB HIV infection with ID in the progression phase without ART and with verified comorbidity of DPT, CVP, and PCP; b - Patient with stage IVB HIV infection with ID in the progression phase without ART and with verified comorbidity of DPT and PCP.

**Fig. 2.**
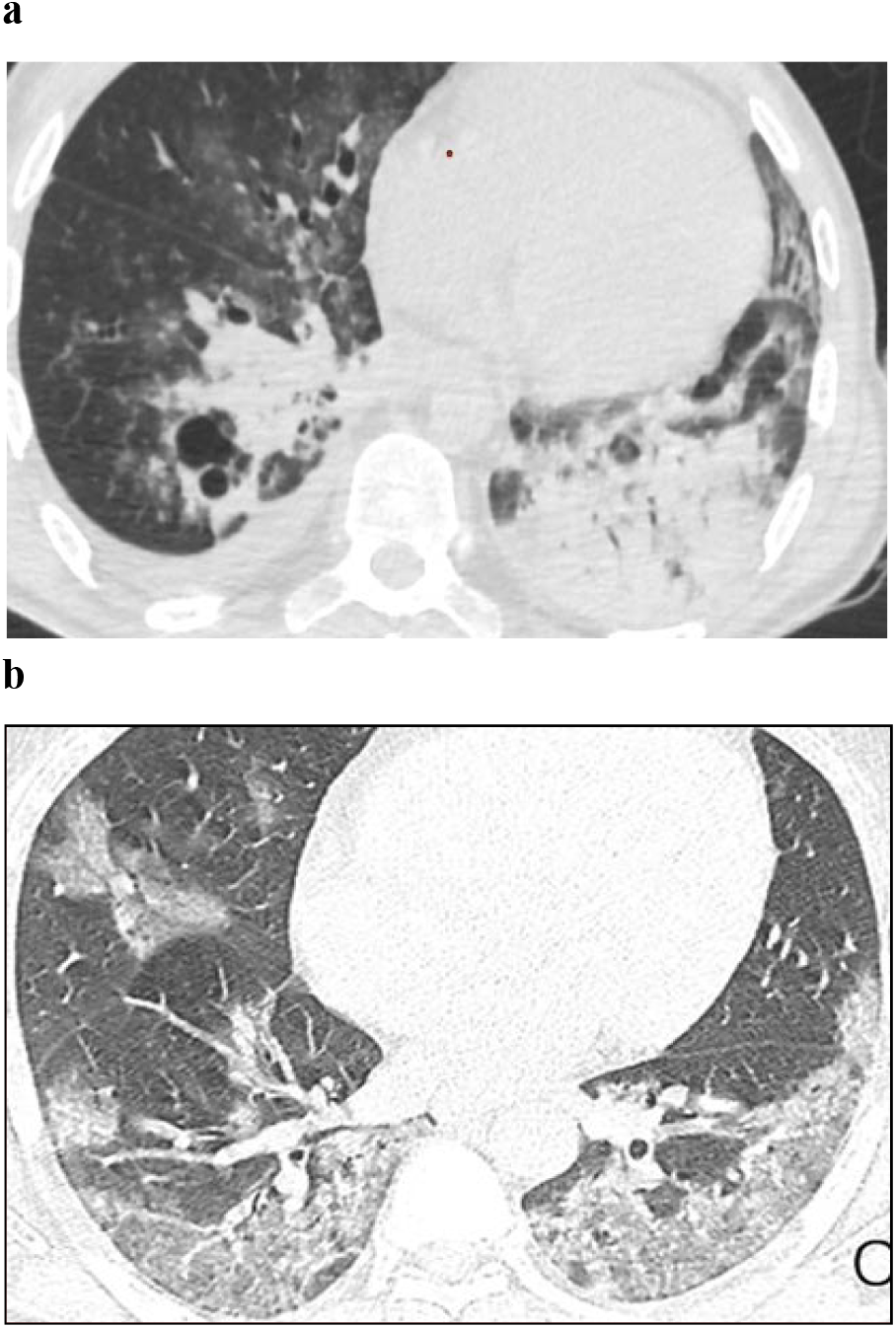
The CT of the ChO. Axial projection, pulmonary window mode: a - Patient with IVB stage HIV infection with ID in the progression phase without ART and with verified comorbidity of DPT, CVP, and PCcP; b - Patient with stage IVB HIV infection with ID in the progression phase without ART and with verified comorbidity of DPT and PCcP.

According to Figures 1 and 2, four syndromes of the same type are visualized on CT of the ChO: dissemination, pleural pathology, enhancement and deformation of the pulmonary pattern with “ground glass” areas and adenopathy.

Thus, on CT of the ChO in patients with comorbidity of tuberculosis of the respiratory system (TRS), CVP and PCP or TRS and PCP and without CVP in IVB stage of HIV infection in the progression phase and the absence of ART with ID, the same type of overlapping syndromes of several pathologies were visualized. It was not possible to differentiate them from each other, because it requires timely diagnosis using microbiological, molecular genetics and immunological studies to detect *M. tuberculosis, SARS-CoV-*2, *P. jirovecii, S. pneumoniae*, which is necessary for adequate timely etiological treatment.

The distribution of patients in the observed groups by frequency and nature of drug resistance *of M. tuberculosis* to ATD is presented **in Table 2**.

**Table 2.** Distribution of patients in groups by frequency and nature of drug resistance of *M. tuberculosis* to ATD (M±m)

|  |  |  |  |  |  |  |
| --- | --- | --- | --- | --- | --- | --- |
| Surveillance Team | Number of patients, abs. (%) | Drug susceptibility of *M. tuberculosis* |  |  |  |  |
|  |  | Sensitivity kept | MR | PR | MDR | BDR |
| 1A | 29 (100) | – | – | 6 [20,7±7,5] | 13 [44,8±9,2] | 10 [34,5±8,8] |
| 2A | 31 (100) | – | – | 7 [22,6±7,5] | 17 [54,8±8,9] | 7 [22,6±7,5] |
| 1B | 29 (100) | – | – | 7 [24,1±7,9] | 14 [48,3±9,2] | 8 [27,6±8,3] |
| 2B | 31 (100) | – | 2 [6,4±4,4] | 8 [25,8±7,8] | 14 [45,2±8,9] | 7 [22,6±7,5] |

As follows from Table 2, *M. tuberculosis* had polyresistance (PR) to two ATDs, but not to combination of isoniazid and rifampicin: in group 1A - 20.7% of patients, in group 2A - 22.6%, in group 1B - 24.1% and in group 2B - 25.8% (*p*> 0.05); monoresistance (MR) was noted in only 2 (6.4%) patients, while patients susceptible to all ATDs were not revealed. In greatest number of patients from the observed groups, *M. tuberculosi*s had multidrug resistance (MDR) to the combination of isoniazid and rifampicin and broad drug resistance (BDR) to the combination of isoniazid, rifampicin, fluoroquinolones, and injectable aminoglycosides (streptomycin, kanamycin and amikacin) and capreomycin. MDR in group 1A was detected in 44.8% of cases, in group 2A - in 54.8%, in group 1B - in 48.3% and in group 2B - in 45.2% (p> 0.05), and BDR, respectively, in 34.5, 22.6, 27.6 and 22.6% of cases (p> 0.05).

Previously, we have described high levels of MDR and BDR *of M. tuberculosis* in patients with caseous pneumonia with common caseous necrotic lesions of the lung tissue and severe ID when mean number of CD4+ lymphocytes did not exceed 40 cells per 1 μL of blood. At the same time, the immune system lost control over rampant reproduction of *M*. tuberculosis because frequency of spontaneous and induced mutations increased sharply with formation of drug resistance. Similar mutations occur in natural “wild” strains even before contact with ATD and develop in the process of inadequate chemotherapy for tuberculosis with formation of MDR and BDR *M. tuberculosis*. A similar mechanism for development of MDR and BDR of *M. tuberculosi*s as we have described also takes place in the late stages of HIV infection with severe ID in patients with tuberculosis and common caseous necrotic lung lesions. In those cases, the pronounced ID in patients with HIV infection was aggravated by comorbidity of OLI tuberculosis including with CVP, PCP and PCcP which causes severe clinical manifestations of the disease and high level of MDR and BDR of *M. tuberculosis* to ATD.

At the same time, we found that in patients with caseous pneumonia with severe ID, the method of sowing material from the respiratory tract revealed in the diagnostic titer bacterial pathogens including *S. pneumoniae* with high level of resistance to broad-spectrum antibiotics (BSA) which significantly aggravated the course of caseous pneumonia and PCcP. In patients from the observed groups, there were multiple resistance of *S. pneumoniae* to beta-lactam antibiotics, tetracyclines, macrolides, levomycetin and other BSA in more than 80% of cases as well as to antibiotics used to treat tuberculosis: rifampicin, levofloxacin, moxifloxacin, amoxicillin + clavulanic acid, clarithromycin and meropenem. Only linezolid proved to be a highly effective drug for treatment of patients with DPT caused by MDR and BDR *M. tuberculosis* and PCcP - *S. pneumoniae* with multiple resistance to BSA.

## Discussion

Comorbidity of DPT with isolation *of M. tuberculosis*, SARS-CoV-2-induced CVP, *P. jirovecii*-caused PCP and *S. pneumoniae*-caused PCcP in the advanced stages of HIV in the progression phase and in the absence of ART was diagnosed, typically 6-9 years after recognition of HIV infection in persons of reproductive age, unemployed, living not in family drug-dependent, alcohol-consuming and tobacco-smoking individuals with concomitant viral hepatitis B or C and COPD. This comorbidity was characterized by pronounced ID (number of CD4+ lymphocytes was less than 50 cells per 1 μL of blood and with the mean number not exceeding 30 cells per 1 μL of blood), high viral load - more than 500 thousand RNA HIV copies in 1 mL of blood), tuberculosis generalization with extrapulmonary lesions and severe CTP, PCP, and PCcP.

At the same time, the clinical picture was characterized by intoxication syndrome, general inflammatory and respiratory manifestations, the pattern was almost the same and nonspecific and features of the clinical course of comorbid disease indicate similarity of the clinical manifestations and course of DPT, CVP, PCP and PCcP which complicates visualization of the CT changes consisting of a complex of four pathological syndromes: dissemination, pleural pathology, enhancing pulmonary pattern and adenopathy. Simultaneous overlap of several pathologies with the same types of clinical manifestations and CT-changes requires a comprehensive etiological diagnosis of the specific diseases. At the same time, in these patients in the late stages of HIV with severe ID, microbiological and molecular genetic studies of diagnostic material from the respiratory system are necessary to determine drug resistance to all drugs that have been used for the treatment of tuberculosis and PCcP, taking into account the fact that M. *tuberculosis* has a high incidence of MDR and BDR - in more than 80% of cases and in *S. pneumoniae* - with multiple resistance an BSA.

## Conclusion

Patients with DPT and HIV infection are at high risk of COVID-19 infection and development of CVP. In cases of severe ID, PCP, and PCcP they should be routinely subjected to preventive comprehensive examinations for timely detection of COVID-19, CVP, PCP, and PCcP as well as of other OLIs for their emergency hospitalization for timely complex treatment and reduction of mortality.

## Data Availability

All data produced in the present work are contained in the manuscript.

## Ethical approval

The study was approved by the local ethics committee of the Yevdokimov Moscow State University of Medicine and Dentistry (former name of the Russian University of Medicine, protocol №677, 7.10.2022). The approval and procedure for the protocol were obtained in accordance with the principles of the Helsinki Convention.

## Consent for publication

Written consent was obtained from the patients for publication of relevant medical information and all of accompanying images within the manuscript.

## Author Credentials

**Mishina Anastasiya** Vladimirovna - Holder of first-level doctoral degree (Kandidat nauk) in medicine, Associate Professor at the Department of Phthisiology and Pulmonology at the FSBEI HE Russian University of Medicine of the Ministry of Health of Russia, Moscow, Russia and the physician of the Department for patients with tuberculosis and HIV infection of FBHI The Tuberculosis Clinical Hospital N 3 named after Professor G.A. Zakharyin of the Ministry of Health of Russia, Moscow, Russia

**Mishin Vladimir** Yur’yevich - Holder of second-level doctoral degree (Doktor nauk) in medicine, Professor, Head of the Department of Phthisiology and Pulmonology at the FSBEI HE Russian University of Medicine of the Ministry of Health of Russia, Moscow, Russia and Consultant Professor at FBHI The Tuberculosis Clinical Hospital N 3 named after Professor G.A. Zakharyin of the Ministry of Health of Russia, Moscow, Russia

**Shashenkov Ivan** Vasil’yevich - Assistant Professor at the Department of Phthisiology and Pulmonology of FSBEI HE FSBEI HE Russian University of Medicine of the Ministry of Health of Russia, Moscow, Russia

